# Age-specific trends in colorectal, appendiceal, and anal tumour incidence by histological subtype in Australia from 1990 to 2020: a population-based time-series analysis

**DOI:** 10.1101/2025.04.21.25326138

**Authors:** Aaron L. Meyers, James G. Dowty, Khalid Mahmood, Finlay A. Macrae, Christophe Rosty, Daniel D. Buchanan, Mark A. Jenkins

**Author notes:** Corresponding author. Centre for Epidemiology and Biostatistics, Melbourne School of Population and Global Health, University of Melbourne, Level 3, 207 Bouverie Street, Carlton, Victoria 3053, Australia.

## Abstract

**Background:** Early-onset bowel cancer incidence (age <50 years) has increased worldwide and is highest in Australia, but how this varies across histology and anatomical site remains unclear. We aimed to investigate appendiceal, proximal colon, distal colon, rectal, and anal cancer incidence trends by age and histology in Australia.

**Methods:** Cancer incidence rate data were obtained from all Australian cancer registries (1990-2020 period). Birth cohort-specific incidence rate ratios (IRRs) and annual percentage change in rates were estimated using age-period-cohort modelling and joinpoint regression.

**Findings:** After excluding neuroendocrine neoplasms, early-onset cancer incidence rose 5-9% annually, yielding 5,341 excess cases (2 per 100,000 person-years; 12% appendix, 45% colon, 36% rectum, 7% anus; 20-214% relative increase). Trends varied by site, period, and age: appendiceal cancer rose from 1990-2020 in 30-49-year-olds; colorectal cancers rose from around 1990-2010 in 20-29-year-olds and from 2010-2020 in 30-39-year-olds; anal cancer rose from 1990-2009 in 40-49-year-olds. Across all sites, IRRs increased with successive birth cohorts since 1960. Notably, adenocarcinoma incidence in the 1990s versus 1950s birth cohort was 2-3-fold for colorectum and 7-fold for appendix. The greatest subtype-specific increases occurred for appendiceal mucinous adenocarcinoma, colorectal non-mucinous adenocarcinoma, and anal squamous cell carcinoma. Only later-onset (age ≥50) colorectal and anal adenocarcinoma rates declined. Appendiceal tumours, neuroendocrine neoplasms (all sites), anorectal squamous cell carcinomas, and colon signet ring cell carcinomas rose across early-onset and later-onset strata.

**Interpretation:** Appendiceal, colorectal, and anal cancer incidence is rising in Australia with variation across age and histology, underscoring the need to identify factors driving these trends.

**Funding:** ALM is supported by an Australian Government Research Training Program Scholarship, Rowden White Scholarship, and WP Greene Scholarship. DDB is supported by a National Health and Medical Research Council of Australia (NHMRC) Investigator grant (GNT1194896), a University of Melbourne Dame Kate Campbell Fellowship, and by funding awarded to The Colon Cancer Family Registry (CCFR, www.coloncfr.org) from the National Cancer Institute (NCI), National Institutes of Health (NIH) [award U01 CA167551]. MAJ is supported by an NHMRC Investigator grant (GNT1195099), a University of Melbourne Dame Kate Campbell Fellowship, and by funding awarded to the CCFR from NCI, NIH [award U01 CA167551].

## Introduction

Colorectal cancer is the third most common cancer and second leading cause of cancer-related deaths worldwide, accounting for almost 2 million new cases and 1 million deaths in 2022 (1). The incidence of early-onset colorectal cancer (diagnosed before age 50 years) is increasing across several high-income countries, with rates now highest in Australia (2-4). This contrasts stabilising or declining incidence among Australians above age 50, a phenomenon attributed to population-based screening available to people aged 50-74 (2-4). The cause of increasing early-onset incidence is unknown, although increasing risk factor exposure among younger generations is hypothesised (2-4). This is supported by reports of a birth cohort effect, with colorectal cancer incidence 3-4-fold higher among Australians born in the 1980s versus 1950s (4). Risk factors that have increased in prevalence in Australia include alcohol abuse (5), metabolic syndrome, obesity, insufficient fruit (6-8), physical inactivity (9), and diabetes (10) – all of which are associated with early-onset colorectal cancer (11). Nevertheless, establishing drivers of the growing disease burden remains elusive, complicated by the aetiological variability of colorectal cancer.

Colorectal cancers encompass a range of subtypes with extensive risk factor heterogeneity. Diabetes more strongly associates with proximal (right-sided) than distal (left-sided) colorectal cancer (12). Conversely, physical activity and obesity more strongly associate with distal and proximal colon cancer, while aspirin and smoking more strongly associate with rectal cancer (13,14). These patterns could be due to risk factor associations varying by clinicopathological features. For example, proximal colon cancers are more likely to develop from serrated polyps and harbour mucinous and signet ring cell morphology (15,16). Tumour pathology can also reflect aetiology that varies by age, with early-onset colorectal cancers preferentially localising to the distal colorectum and displaying aggressive histology, including poor differentiation and mucinous or signet ring differentiation, compared with later-onset cases (3,17). Such differences suggest that risk factors may differ for early-onset disease.

Few studies have examined which subtypes drive the rising incidence, with inconsistent findings. In Europe and Australia, the rise appears more pronounced for colon than rectal cancer, while the opposite is reported for North America (2,18,19). This could reflect inconsistent classification, with appendix defined as colon cancer by European and Australian but not North American registries (2-4,18,19). An alternative explanation is an artefactual increase from reclassifying low-grade neuroendocrine neoplasms as malignant in early 2000s, which could upwardly bias the rectal-to-colon cancer ratio that is reportedly characteristic of early-onset disease (19,20). This underscores the importance of analysing colorectal cancer trends by anatomical and histological subtype.

Anal cancer incidence has also increased, particularly among adults aged <60 years (21), suggesting that causes for rising early-onset cancer may affect risk across the lower gastrointestinal tract. However, no study has explored temporal dynamics of appendiceal, colorectal, and anal cancer histotypes in Australia. Such an analysis is crucial to highlight shared or distinct risk factors driving recent trends, identify subtypes warranting further aetiological investigation, and inform discourse around lowering the age for population screening, which currently targets colorectal adenocarcinoma. To address these gaps, we present for the first time an analysis of variations in appendiceal, proximal colon, distal colon, rectal, and anal cancer incidence by histological subtype in Australia across age, period, and birth cohort.

## Methods

### Study design and data sources

This retrospective, nationwide time-series included data on all incident appendiceal, colorectal, and anal cancers diagnosed in Australia from 1990-2020. Cases were defined by anatomical site of the primary diagnosis. Aggregated cancer records for each histology, topography, sex, diagnosis year, and 10-year age group (20-29 to 90+ years) were obtained from the Australian Institute of Health and Welfare, which collates population-based data from all state and territory cancer registries (22). Corresponding population figures by sex, age, and year were obtained from the Australian Bureau of Statistics (23).

### Tumour subtyping

The following malignant subtype categories were included: appendix (International Classification of Diseases for Oncology, 3^rd^ revision: C18.1), proximal colon (cecum, ascending colon, hepatic flexure, transverse colon, splenic flexure; C18.0, C18.2-C18.5), distal colon (descending and sigmoid colon; C18.6, C18.7), overlapping/unspecified colon (C18.8, C18.9), rectum (C19.9, C20.9), and anus (C21). Tumours were further classified into non-overlapping histological subtypes: adenocarcinoma not otherwise specified (NOS); adenocarcinoma in a polyp; mucinous adenocarcinoma; signet ring cell carcinoma; squamous cell carcinoma; carcinoma NOS; neuroendocrine neoplasm; melanoma; sarcoma; other (all remaining codes) (Appendix Table S1).

### Statistical analysis

The annual prevalence of histotypes by sex, anatomical site, and age (20-49, ≥50 years) was summarised using direct age-standardisation to the median (2005) case population. We selected age strata used in previous studies (2,11,17) to capture changing histotype distributions between early-onset and later-onset cases.

Annual age-specific and site-specific tumour incidence rates (IRs) per 100,000 person-years were estimated overall and by sex and histology. To quantify overall burden, we combined tumours across histological subtypes but excluded neuroendocrine neoplasms to avoid confounding from reclassifying indolent subtypes to malignant across the early 2000s, as previously demonstrated (2,19,20). Secondary analyses were undertaken with neuroendocrine neoplasms included to assess the impact on results (2,19,20).

For evaluating period effects, joinpoint regression models were fitted to age-specific cancer rates to identify inflection years (joinpoints) for changes in the direction and magnitude of trends (24,25). Model selection for the optimal joinpoint number was based on a weighted Bayesian information criterion (24,25). The magnitude of rate changes was expressed as annual percentage change (APC), quantified using the gradient of each segmented period. Average annual percentage change (AAPC) was based on a weighted geometric mean of APC across segments. Confidence intervals (CIs) were computed using the empirical quantile method with 5,000 resamples. We assumed uncorrelated errors and homoscedasticity. To obtain stable estimates, we truncated periods with 0 cases and limited subgroup analyses to common histotypes (prevalence >3%) (Appendix pp 5-11).

To evaluate birth cohort trends, independent of age and period, we fitted age-period-cohort models to each tumour subtype using weighted least squares, assuming Poisson-distributed counts and overdispersion parameters for excess-Poisson variation (26). Ten-year intervals were used to define eight age groups (20-29 to 90-99), three diagnosis periods (1990-1999, 2000-2009, 2010-2019), and 10 birth cohorts for each decade from 1900-1999. We computed incidence rate ratios (IRRs) comparing net IR in each cohort to the 1950 (middle) cohort. AAPC in rates between 1990 and 2019 was obtained from model-based estimates for each age stratum (local drifts) using the gradient of IRR curves. Wald tests were used to assess non-linear components and identify heterogeneity of local drifts compared to age-standardised AAPC (net drift).

Excess early-onset cases attributable to rising incidence were estimated per annum in 10-year age groups as the difference between observed and expected numbers without rate increases from 1990-2020. Age-specific AAPCs from joinpoint modelling were decomposed into absolute attributable cases using corresponding population numbers. Men, women, and colon subsites were combined because they had comparable rates and trends (see results). In 40-49-year-olds, we modelled excess colon cancers from 2003 given strong prior incidence decreases (*p*<0·000001). P-scores were defined as the ratio of excess to expected cases, expressed as a percentage, where negative values correspond to fewer cases than expected and positive values correspond to increasing relative excess cases (27). Absolute excess risk (AER) was defined as the number of excess cases per 100,000 person-years. Uncertainty intervals (UI) were derived from the CI of the AAPC. All hypothesis tests were two-sided and tested at alpha = 0·05. Statistical analyses were undertaken using Joinpoint Regression Program software version 5.2 (28) and R version 4.2.2 (29).

## Results

Between 1990 and 2020, a total of 413,691 colorectal, appendiceal, and anal tumours were diagnosed among Australians aged ≥20 years (appendix, 1·9%; proximal colon, 35·2%; distal colon, 22·5%; overlapping/unspecified colon, 5·3%; rectum, 32·6%; anus, 2·4%). Of these, 34,907 (8·4%) occurred before age 50 (early-onset), which were evenly distributed between sexes (50·1% men). Most early-onset tumours were localised to the rectum (38·3%), followed by proximal colon (22·8%), distal colon (22·6%), appendix (10·8%), and anus (4·0%). After excluding neuroendocrine neoplasms, IRs were highest for rectal cancer among 20-69-year-olds, whereas proximal colon cancer rates were highest in those aged ≥70 (Fig. 1). Trends were consistent between men and women (Appendix Fig. S2 and S3).

**Figure 1:**
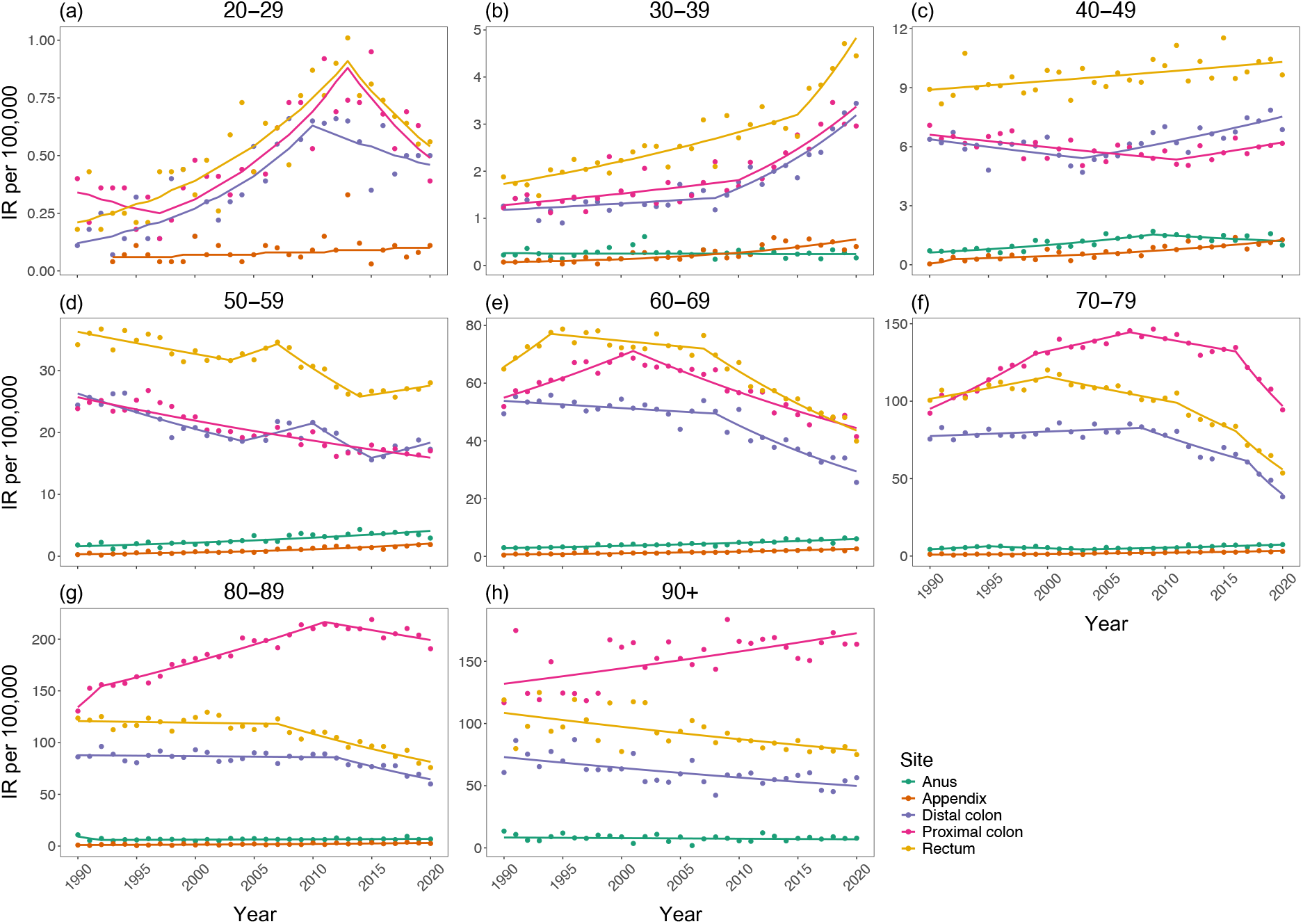
Period trends in cancer incidence by anatomical site and age from 1990 to 2020. Joinpoint regression was used to fit a series of joined straight lines to cancer rates per 100,000 Australians aged **a:** 20-29; **b:** 30-39; **c:** 40-49; **d:** 50-59; **e:** 60-69; **f:** 70-79; **g:** 80-89; and **h:** 90+ years. The optimal number of segments was based on the weighted Bayesian information criterion method. Annual percentage changes and 95% confidence intervals for each segment are given in Table 1. Data exclude neuroendocrine neoplasms. Results for all histological subtypes combined and by sex and histology are presented in the Appendix pp 2-4 and 13-23. IR, incidence rate.

Since 1990, multiple periods of increasing early-onset proximal and distal colon cancer were detected after excluding neuroendocrine neoplasms (Fig. 1, Table 1). For ages 20-29 years, increases occurred from 1997-2013 for proximal colon (APC 8·3% [95% CI: 5·8-28·9]) and from 1990-2010 for distal colon cancer (8·6% [6·1-20·5]). The rise began over a decade later for 30-39-year-olds, with a 6·4% (95% CI: 2·8-24·6) per annum increase from 2010 for proximal colon and 6·9% (4·4-15·0) per annum increase from 2008 for distal colon cancer. A modest increase in distal colon cancer was detected among 40-49-year-olds since 2003 (APC 2·0% [95% CI: 1·1-4·2]), but no increases were observed for proximal colon cancer. Among those aged ≥50 years, multiple periods of decreasing incidence were observed, with variable induction times. Decreases were larger for distal than proximal colon cancer. Since 1990, the AAPC in early-onset colon cancer rates across subsites (including unknown/overlapping) ranged from 1·2-3·5% (Table 2). This resulted in 2,398 (95% UI: 1,415-3,557) excess cases, equivalent to 1 per 100,000 person-years or a 25·5% increase in observed cases compared to expected numbers under no rate increase. Results were similar when including neuroendocrine neoplasms (Table 2, Appendix Table S2).

**Table 1:** Temporal trends in tumour incidence rates by age, anatomical site, and calendar period of diagnosis.

| Age, years | n | Trend 1 |  | Trend 2 |  | Trend 3 |  | Trend 4 |  |
| --- | --- | --- | --- | --- | --- | --- | --- | --- | --- |
|  |  | Period | APC (95% CI) | Period | APC (95% CI) | Period | APC (95% CI) | Period | APC (95% CI) |
| Appendix |  |  |  |  |  |  |  |  |  |
| 20-29 | 81 | 1993-2020 | 2.2 (-0.4, 4.8) | — | — | — | — | — | — |
| 30-39 | 244 | 1990-2020 | 7.4 (5.1, 9.7) | — | — | — | — | — | — |
| 40-49 | 611 | 1990-1992 | 123.7 (19.3, 270.4) | 1992-2020 | 5.4 (2.9, 7.0) | — | — | — | — |
| 50-59 | 798 | 1990-2020 | 6.3 (5.1, 7.5) | — | — | — | — | — | — |
| 60-69 | 910 | 1990-2020 | 4.7 (3.8, 5.6) | — | — | — | — | — | — |
| 70-79 | 788 | 1990-2020 | 4.5 (3.5, 5.5) | — | — | — | — | — | — |
| 80-89 | 368 | 1990-2020 | 3.9 (2.0, 5.9) | — | — | — | — | — | — |
| 90+ | 50 | — | — | — | — | — | — | — | — |
| Proximal colon |  |  |  |  |  |  |  |  |  |
| 20-29 | 482 | 1990-1997 | -4.6 (-24.7, 3.3) | 1997-2013 | 8.3 (5.8, 28.9) | 2013-2020 | -8.0 (-21.6, 0.0) | — | — |
| 30-39 | 1,849 | 1990-2010 | 1.8 (-6.6, 3.3) | 2010-2020 | 6.4 (2.8, 24.6) | — | — | — | — |
| 40-49 | 5,314 | 1990-2011 | -1.0 (-3.4, -0.5) | 2011-2020 | 1.7 (-0.2, 10.0) | — | — | — | — |
| 50-59 | 14,983 | 1990-2020 | -1.6 (-1.9, -1.3) | — | — | — | — | — | — |
| 60-69 | 32,936 | 1990-2001 | 2.4 (1.5, 3.5) | 2001-2020 | -2.4 (-2.9, -2.1) | — | — | — | — |
| 70-79 | 48,249 | 1990-1999 | 3.6 (2.7, 6.2) | 1999-2007 | 1.3 (-0.3, 2.9) | 2007-2016 | -1.0 (-2.7, 0.1) | 2016-2020 | -7.5 (-10.6, -5.6) |
| 80-89 | 35,015 | 1990-1992 | 7.3 (1.9, 12.1) | 1992-2011 | 1.8 (-3.0, 2.1) | 2011-2020 | -0.9 (-2.6, 1.4) | — | — |
| 90+ | 5,311 | 1990-2020 | 0.9 (0.4, 1.4) | — | — | — | — | — | — |
| Distal colon |  |  |  |  |  |  |  |  |  |
| 20-29 | 384 | 1990-2010 | 8.6 (6.1, 20.5) | 2010-2020 | -3.1 (-25.3, 3.7) | — | — | — | — |
| 30-39 | 1,657 | 1990-2008 | 1.1 (-2.1, 2.4) | 2008-2020 | 6.9 (4.4, 15.0) | — | — | — | — |
| 40-49 | 5,633 | 1990-2003 | -1.3 (-4.7, 0.0) | 2003-2020 | 2.0 (1.1, 4.2) | — | — | — | — |
| 50-59 | 15,019 | 1990-2004 | -2.4 (-3.4, -1.8) | 2004-2010 | 2.4 (0.2, 7.6) | 2010-2015 | -5.8 (-10.8, -3.1) | 2015-2020 | 2.9 (0.2, 9.3) |
| 60-69 | 25,644 | 1990-2008 | -0.5 (-1.0, 0.2) | 2008-2020 | -4.2 (-5.6, -3.3) | — | — | — | — |
| 70-79 | 27,593 | 1990-2008 | 0.4 (0.0, 0.9) | 2008-2017 | -3.3 (-4.7, -1.8) | 2017-2020 | -13.4 (-19.6, -9.2) | — | — |
| 80-89 | 14,858 | 1990-2012 | -0.1 (-0.5, 0.5) | 2012-2020 | -3.5 (-7.1, -1.9) | — | — | — | — |
| 90+ | 1,971 | 1990-2020 | -1.3 (-1.8, -0.7) | — | — | — | — | — | — |
| Rectum |  |  |  |  |  |  |  |  |  |
| 20-29 | 519 | 1990-2013 | 6.7 (5.2, 9.4) | 2013-2020 | -7.2 (-25.6, 0.7) | — | — | — | — |
| 30-39 | 2,618 | 1990-2015 | 2.5 (-0.1, 3.2) | 2015-2020 | 8.6 (3.1, 23.8) | — | — | — | — |
| 40-49 | 8,686 | 1990-2020 | 0.5 (0.2, 0.8) | — | — | — | — | — | — |
| 50-59 | 23,261 | 1990-2003 | -1.0 (-2.6, -0.6) | 2003-2007 | 2.0 (-0.4, 4.5) | 2007-2014 | -4.0 (-7.3, -2.7) | 2014-2020 | 1.1 (-0.4, 4.3) |
| 60-69 | 36,621 | 1990-1994 | 4.1 (1.0, 10.2) | 1994-2007 | -0.5 (-1.7, 0.1) | 2007-2020 | -3.8 (-4.6, -3.2) | — | — |
| 70-79 | 36,670 | 1990-2000 | 1.3 (-2.2, 5.3) | 2000-2011 | -1.4 (-4.5, 4.8) | 2011-2016 | -4.0 (-8.8, 1.0) | 2016-2020 | -8.8 (-13.3, -5.3) |
| 80-89 | 19,414 | 1990-2007 | -0.1 (-0.7, 0.6) | 2007-2020 | -2.8 (-4.0, -2.1) | — | — | — | — |
| 90+ | 3,020 | 1990-2020 | -1.1 (-1.6, -0.6) | — | — | — | — | — | — |
| **Anus** |  |  |  |  |  |  |  |  |  |
| 20-29 | 36 | — | — | — | — | — | — | — | — |
| 30-39 | 251 | 1990-2020 | -0.2 (-1.8, 1.4) | — | — | — | — | — | — |
| 40-49 | 1,055 | 1990-2009 | 4.8 (3.5, 7.2) | 2009-2020 | -2.2 (-7.4, 0.6) | — | — | — | — |
| 50-59 | 2,153 | 1990-2020 | 3.2 (2.3, 4.0) | — | — | — | — | — | — |
| 60-69 | 2,574 | 1990-2020 | 2.5 (2.1, 3.0) | — | — | — | — | — | — |
| 70-79 | 2,186 | 1990-1995 | 7.6 (0.3, 22.6) | 1995-2003 | -4.6 (-15.3, -0.4) | 2003-2020 | 3.2 (1.7, 5.7) | — | — |
| 80-89 | 1,198 | 1990-1992 | -19.9 (-30.0, 0.0) | 1992-2020 | 0.6 (-0.2, 3.1) | — | — | — | — |
| 90+ | 267 | 1990-2020 | -0.6 (-2.5, 1.3) | — | — | — | — | — | — |

Note: Excludes neuroendocrine neoplasms. Estimates for all histotypes are provided in Table S2.
APC, annual percentage change; CI, confidence interval; *n*, number of cases.

**Table 2:** Cumulative excess early-onset tumours attributable to rising incidence rates between 1990 and 2020.

| Age | Excluding neuroendocrine neoplasms |  |  |  |  | All histological subtypes |  |  |  |  |
| --- | --- | --- | --- | --- | --- | --- | --- | --- | --- | --- |
|  | AAPC (95% CI) | Recorded cases | Excess cases (95% UI) | AER (95% UI)<sup>a</sup> | P-score (%; 95% UI) | AAPC (95% CI) | Recorded cases | Excess cases (95% UI) | AER (95% UI)<sup>a</sup> | P-score (%; 95% UI) |
| **Appendix** |  |  |  |  |  |  |  |  |  |  |
| 20-29 | 4.4 (-0.3, 9.5) | 81 | 32 (-1, 81) | 0.0 (0.0, 0.1) | 66.1 (-1.8, Inf) | 5.5 (4.3, 6.7) | 1,290 | 753 (515, 1,078) | 0.8 (0.6, 1.2) | 160.2 (109.6, 229.2) |
| 30-39 | 7.4 (5.1, 9.7) | 244 | 181 (95, 244) | 0.2 (0.1, 0.3) | 290.5 (63.9, Inf) | 6.2 (5.1, 7.3) | 1,123 | 712 (515, 969) | 0.7 (0.5, 1.0) | 196.6 (142.2, 267.6) |
| 40-49 | 6.8 (4.7, 9.1) | 611 | 424 (222, 611) | 0.5 (0.2, 0.7) | 226.4 (57.2, Inf) | 6.6 (5.5, 7.8) | 1,291 | 883 (637, 1,209) | 1.0 (0.7, 1.3) | 223.0 (160.9, 305.3) |
| Total | — | 936 | 638 (316, 936) | 0.2 (0.1, 0.3) | 213.6 (51.0, Inf) | — | 3,704 | 2,348 (1,667, 3,256) | 0.8 (0.6, 1.2) | 191.2 (135.7, 265.0) |
| **Colon** |  |  |  |  |  |  |  |  |  |  |
| 20-29 | 3.5 (2.4, 4.7) | 939 | 415 (243, 646) | 0.4 (0.3, 0.7) | 79.2 (34.9, 220.2) | 3.6 (2.4, 4.8) | 958 | 434 (258, 675) | 0.5 (0.3, 0.7) | 83.9 (49.8, 130.4) |
| 30-39 | 2·6 (1·9, 3·3) | 3,725 | 1,262 (844, 1,761) | 1·3 (0·9, 1·8) | 51·3 (29·3, 89·7) | 2·6 (2·0, 3·4) | 3,808 | 1,327 (910, 1,830) | 1·4 (1·0, 1·9) | 55·0 (37·7, 75·9) |
| 40-49<sup>b</sup> | 1·2 (0·6, 1·9) | 7,158 | 721 (328, 1,150) | 1·3 (0·6, 2·0) | 11·2 (4·8, 19·1) | 1·0 (0·5, 1·9) | 7,285 | 639 (274, 1,239) | 1·1 (0·5, 2·2) | 9·6 (4·1, 18·6) |
| Total | – | 11,822 | 2,398 (1,415, 3,557) | 1·0 (0·6, 1·4) | 25·5 (13·6, 43·0) | – | 12,051 | 2,400 (1,442, 3,744) | 1·0 (0·6, 1·5) | 25·0 (15·0, 39·0) |
| **Rectum** |  |  |  |  |  |  |  |  |  |  |
| 20-29 | 4·5 (2·9, 6·2) | 519 | 275 (150, 466) | 0·3 (0·2, 0·5) | 113·0 (40·5, 878·4) | 5·5 (4·1, 7·0) | 622 | 376 (237, 580) | 0·4 (0·2, 0·6) | 159·8 (100·7, 246·5) |
| 30-39 | 3·0 (2·5, 3·5) | 2,618 | 1,013 (796, 1,260) | 1·1 (0·8, 1·3) | 63·1 (43·7, 92·8) | 3·5 (3·0, 3·9) | 3,027 | 1,326 (1,096, 1,584) | 1·4 (1·1, 1·7) | 79·1 (65·4, 94·4) |
| 40-49 | 0·5 (0·2, 0·8) | 8,686 | 661 (231, 1,159) | 0·7 (0·3, 1·3) | 8·2 (2·7, 15·4) | 0·9 (0·6, 1·2) | 9,470 | 1,308 (840, 1,839) | 1·5 (0·9, 2·0) | 16·1 (10·3, 22·6) |
| Total | – | 11,823 | 1,949 (1,177, 2,885) | 0·7 (0·4, 1·0) | 19·7 (11·1, 32·3) | – | 13,119 | 3,010 (2,173, 4,003) | 1·1 (0·8, 1·4) | 29·9 (21·6, 39·8) |
| **Anus** |  |  |  |  |  |  |  |  |  |  |
| 20-29 | – | 36 | – | – | – | – | 36 | – | – | – |
| 30-39 | -0·2 (-1·8, 1·4) | 251 | -8 (-58, 65) | 0·0 (-0·1, 0·1) | -3·2 (-18·8, 34·9) | -0·1 (-1·7, 1·5) | 258 | -4 (-55, 70) | 0·0 (-0·1, 0·1) | -1·6 (-22·2, 28·3) |
| 40-49 | 2·6 (1·6, 3·6) | 1,055 | 364 (203, 570) | 0·4 (0·2, 0·6) | 52·7 (23·9, 117·6) | 2·7 (1·7, 3·8) | 1,075 | 389 (220, 606) | 0·4 (0·2, 0·7) | 57·6 (32·5, 89·8) |
| Total | – | 1,342 | 356 (145, 635) | 0·2 (0·1, 0·3) | 37·5 (12·5, 94·6) | – | 1,369 | 385 (165, 676) | 0·2 (0·1, 0·4) | 41·7 (17·8, 73·3) |

CI, confidence interval; AAPC, average annual percentage change; UI, uncertainty interval; AER, absolute excess risk.
<sup>a</sup>Per 100,000 persons-years.
<sup>b</sup>2003-2020 period.

For rectal cancer (excluding neuroendocrine neoplasms), IRs increased per annum among 20-29-year-olds until 2013 (APC 6·7% [95% CI: 5·2-9·4]) (Fig. 1, Table 1). Similar increases were detected in 30-39-year-olds from 2015. A moderate linear increase was observed among 40-49-year-olds since 1990 (APC 0·5% [95% CI: 0·2-0·8]). Conversely, rates generally decreased over the past decade by approximately 4% per annum among those aged ≥50 years. Since 1990, the AAPC in early-onset rectal cancer rates ranged from 0·5-4·5% (Table 2). This resulted in 1,949 (95% UI: 1,177-2,885) excess cases, equivalent to 0·7 per 100,000 person-years or a 19·7% relative increase. Results were inflated when including neuroendocrine neoplasms (Table 2, Appendix Table S2).

Appendiceal cancer (excluding neuroendocrine neoplasms) displayed linear increases for all age groups except 20-29-year-olds, where increases were only observed for neuroendocrine neoplasms (Table 2, Appendix Tables S2 and S4). Since 1990, the AAPC in early-onset appendiceal tumour rates ranged from 5·5-6·6% (Table 2). Trends were similar after excluding neuroendocrine neoplasms (75% of early-onset cases). Increasing early-onset IRs resulted in 2,348 (95% UI: 1,667-3,256) and 638 (316-936) excess tumours with and without including neuroendocrine neoplasms, equivalent to 0·8 and 0·2 cases per 100,000 person-years or a 191·2% and 213·6% relative increase, respectively.

Anal cancer incidence (excluding neuroendocrine neoplasms) increased for all age groups 40-79 years, but no trends were detected among other ages (Fig. 1, Table 1). Since 1990, the AAPC in early-onset anal cancer rates was only remarkable among 40-49-year-olds (2·6% [95% CI: 1·6-3·6]) (Table 2). This resulted in 364 (95% UI: 203-570) excess cases, equivalent to 0·4 per 100,000 person-years (52·7% relative increase).

The age-standardised prevalence of histotypes by tumour site and early-onset (age 20-49 years) versus later-onset diagnosis (age ≥50 years) is presented in Fig. 2. Sex-specific results are given in Appendix Fig. S4 and S5. Histotype heterogeneity by age was most noticeable for appendiceal tumours, with neuroendocrine neoplasms accounting for most early-onset but only about half of later-onset cases. Moreover, mucinous adenocarcinoma accounted for <10% of early-onset but approximately half of later-onset appendiceal cancers. For colorectal tumours, signet ring cell carcinomas and neuroendocrine neoplasms were more prevalent in early-onset cases. For anal cancers, adenocarcinomas comprised a larger share of older-onset cases. Relative histotype distributions were generally consistent, although colorectal neuroendocrine neoplasm prevalence increased over time. Joinpoint trends were consistent when restricting to dominant histotypes, but early-onset colorectal mucinous adenocarcinoma incidence declined or stabilised (Appendix pp 7-11 and 18-23).

**Figure 2:**
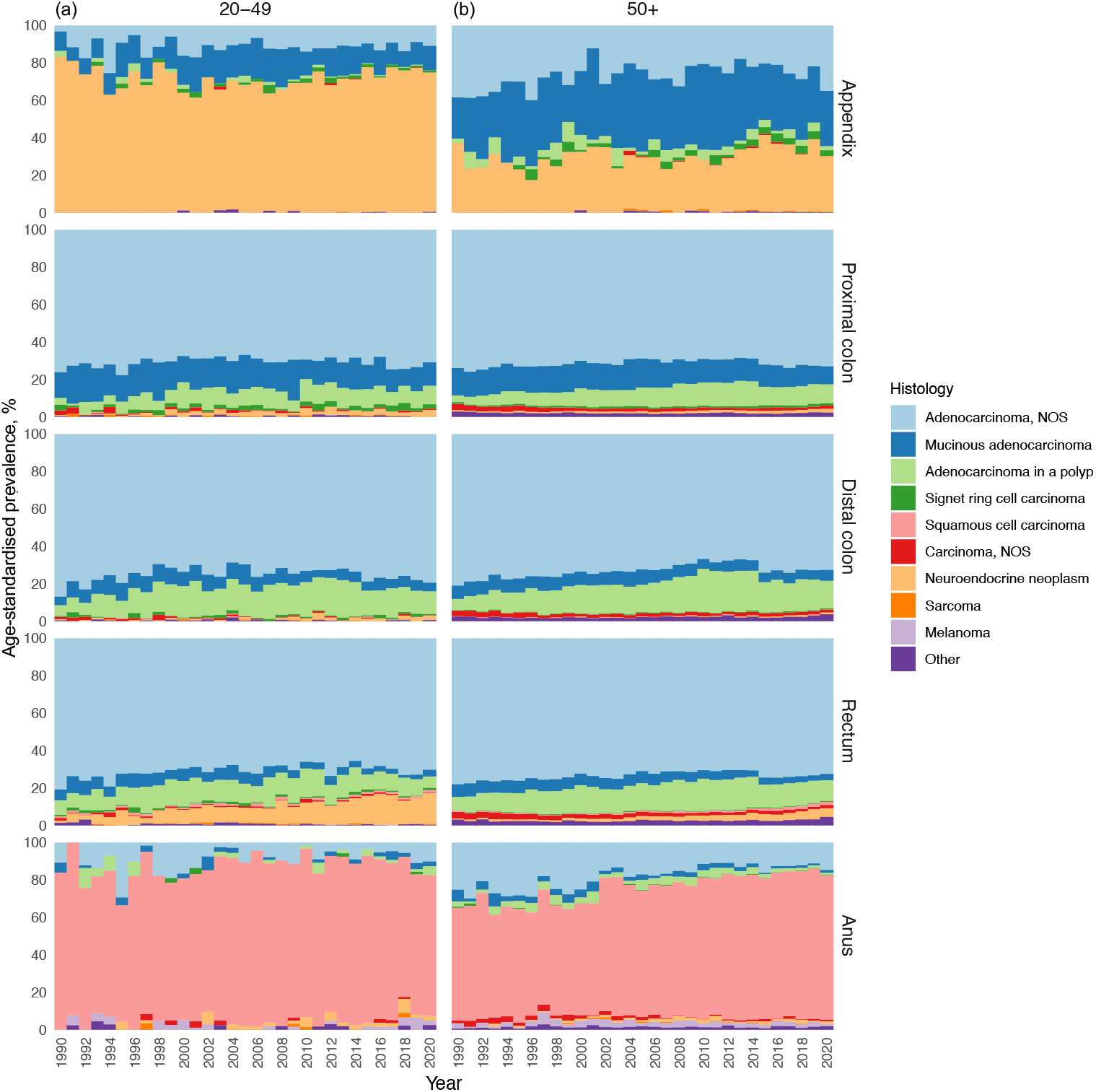
Age-standardised prevalence of tumour histological subtypes by anatomical site and age from 1990 to 2020. Data are presented for **a:** early-onset (age 20-49 years) and **b:** later-onset (age 50+ years) cases, with direct age-standardisation to 2005 case populations. Sex-specific results are given in the Appendix pp 5-6.

Figure 3 summarises birth cohort trends for tumour histotypes. IRRs overall and by sex are given in Appendix pp 12 and 23-24. Compared to those born in the 1950s, IRRs increased with successive birth cohorts for all subtypes except anal adenocarcinoma, colorectal mucinous adenocarcinoma, and rectal signet ring cell carcinoma. Appendiceal tumour subtypes generally exhibited the steepest increase. For adenocarcinoma NOS, IRs in the 1990s compared to 1950s cohort were 6·7-fold for appendiceal and 2-3-fold for colorectal sites. Similar increases were observed for colorectal adenocarcinoma in a polyp. Mucinous adenocarcinoma only increased within the appendix, with 15·9-fold (95% CI: 7·5-34·0) higher rates in the 1990s cohort. For signet ring cell carcinoma, increases were only detected within appendix (IRR 8·2 [95% CI: 1·7-39·1]; 1980 versus 1950) and colon (IRR 2·7 [1·6-4·7]). Squamous cell carcinoma yielded similar, roughly 50% increases since 1960 within rectum and anus. Neuroendocrine neoplasms exhibited the largest increase, and trends were more homogenous across sites compared to other histotypes. In the 1990s compared to 1950s cohort, neuroendocrine neoplasm rates were 12·5-fold (95% CI: 8·6-18·2) for appendiceal, 30·0-fold (17·9-50·3) for rectal, and 6·9-fold (2·4-20·2) for colon cancer. Overall, birth cohort trends were consistent between men and women.

**Figure 3:**
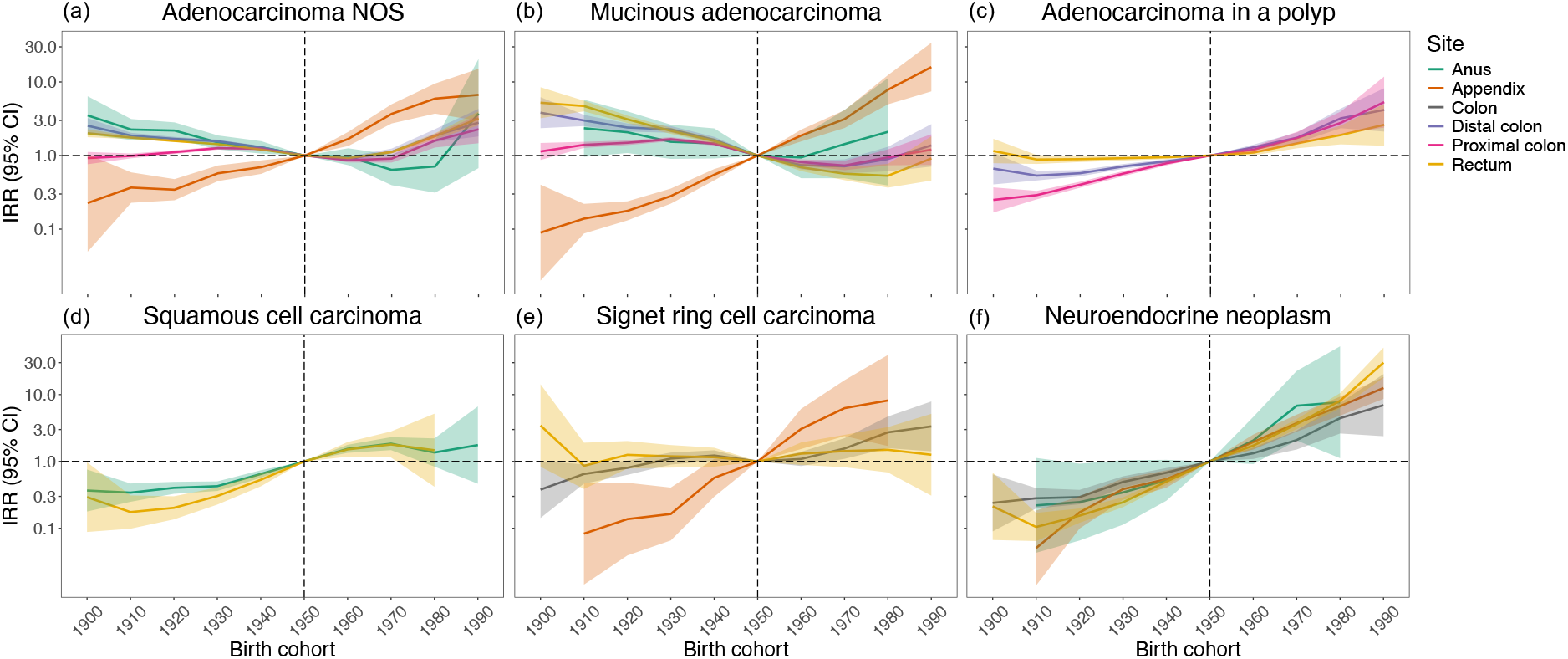
Birth cohort trends in tumour incidence by anatomical site and histology from 1900 to 1990. For each cancer site, age-period-cohort models were fitted to incidence rates from 1990 to 2019 of mutually-exclusive histological subtypes **a:** adenocarcinoma NOS; **b:** mucinous adenocarcinoma; **c:** adenocarcinoma in a polyp; **d:** squamous cell carcinoma; **e:** signet ring cell carcinoma; and **f:** neuroendocrine neoplasm. Shaded areas indicate 95% CIs. The vertical line represents the referent 1950 birth cohort. Neuroendocrine neoplasms and signet ring cell carcinomas of distal, proximal, and overlapping/unspecified colon sites were combined due to small case numbers. Sex-specific results are given in Appendix pp 12 and 23-24. NOS, not otherwise specified; IRR, incidence rate ratio; CI, confidence interval.

To better understand age effects independent of period and birth cohort, we estimated AAPC for histological subtypes from age-period-cohort models, with results presented in Fig. 4 and Appendix pp 12 and 26-28. Trends in increasing incidence were greatest for appendiceal tumour subtypes across all ages (age-standardised AAPC 4-8%). Other subtype-specific trends reflected birth cohort IRRs. Tumours that increased in both young and older age groups and did not conflict with recent joinpoint trends included appendiceal tumours (all histotypes); neuroendocrine neoplasms (all sites); rectal and anal squamous cell carcinoma; and colon signet ring cell carcinoma. For early-onset colorectal adenocarcinoma NOS, trends were similar across subsites and highest among adults aged <40 (AAPC 3-5%). Meanwhile, only colorectal and anal adenocarcinoma NOS rates decreased by around 1-2% per annum for populations older than 50.

**Figure 4:**
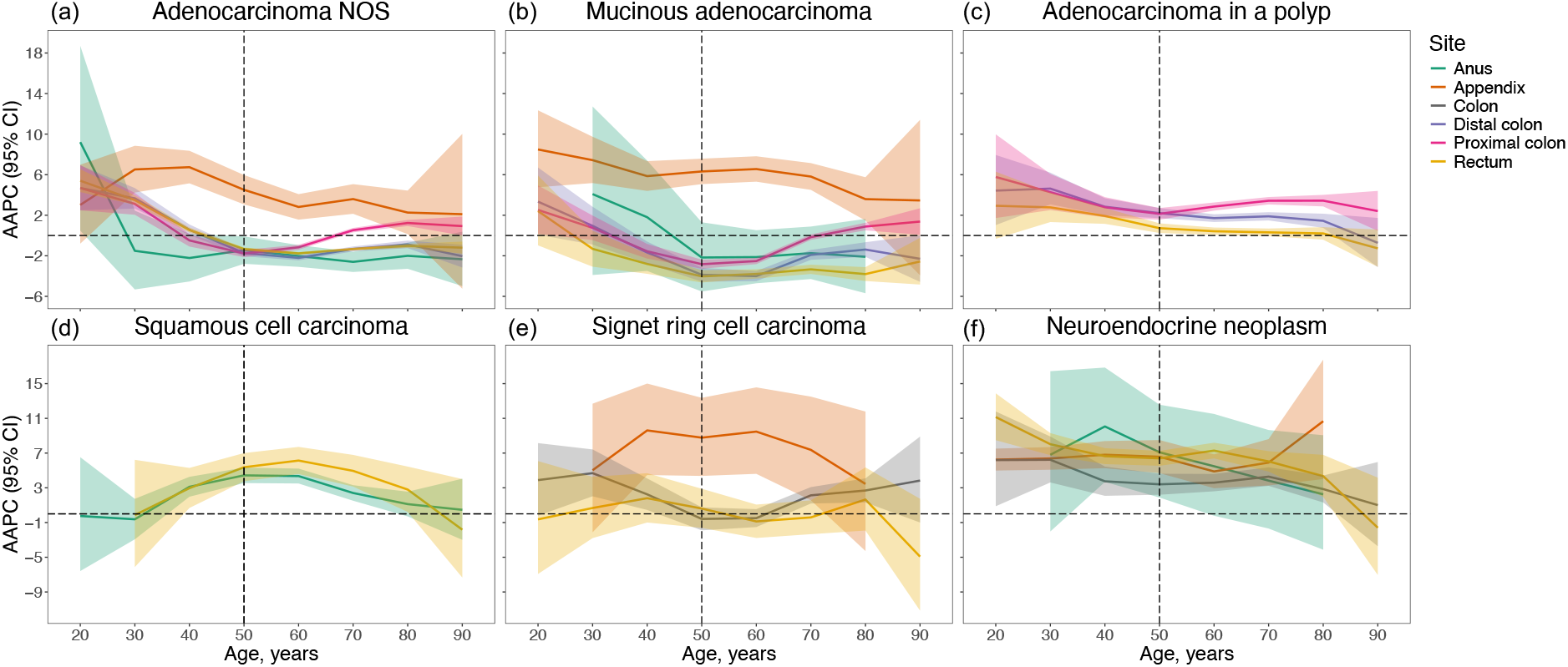
Age-specific AAPC in tumour incidence by anatomical site and histology from 1990 to 2019. For each cancer site, age-period-cohort models were fitted to incidence rates of mutually-exclusive histological subtypes **a:** adenocarcinoma NOS; **b:** mucinous adenocarcinoma; **c:** adenocarcinoma in a polyp; **d:** squamous cell carcinoma; **e:** signet ring cell carcinoma; and **f:** neuroendocrine neoplasm. Shaded areas indicate 95% CIs. The vertical line at 50 years indicates the cut-off for young adults. Neuroendocrine neoplasms and signet ring cell carcinomas of distal, proximal, and overlapping/unspecified colon sites were combined due to small case numbers. Sex-specific results are given in the Appendix pp 12 and 26. NOS, not otherwise specified; AAPC, average annual percentage change; CI, confidence interval.

## Discussion

This population-based time-series offers the most comprehensive and up-to-date estimates of appendiceal, colon, rectal, and anal cancer incidence trends in Australia. While rates of colorectal adenocarcinoma generally declined or stabilised in adults over age 50 years, they continue to rise in younger adults, although this pattern is not observed for other histotypes. Anal and rectal squamous cell carcinomas exhibit similar increases for each 10-year age group from 40-69 years. Our findings of birth cohort effects point toward increasing risks for most evaluated tumour subtypes since 1960. For all age groups, increases were most pronounced for neuroendocrine neoplasms across all sites, and appendiceal tumours irrespective of histology. Despite large relative increases, the absolute impact remains modest, with 2 excess cases per 100,000 person-years from 1990-2020.

The age-related patterning of colorectal cancer trends in this study corroborate previous findings in Australia (2,4). We further demonstrated that divergent trends between early-onset and later-onset cases are specific to conventional adenocarcinoma (adenocarcinoma NOS; non-mucinous, non-signet ring cell). Similar patterns are reported in North America (30-32), Germany (33), and Norway (3), where conventional adenocarcinoma is the only histotype to show increasing incidence among adults under 50, alongside concomitant reductions in older adults since early 2000s. Such results could inform debate around lowering the screening age, as adenocarcinomas arise from precancerous polyps through well-described natural history and are the target of population screening, introduced for ages 50-74 in Australia and other high-income countries in early 2000s (2-4). This likely explains declining rates among older adults, but it remains unclear whether early-onset adenocarcinomas represent a continuum of older-onset disease driven by earlier risk factor exposure.

Increasing rates of adenocarcinoma in a polyp among young, but not older, adults may indicate that factors behind the rise shorten the polyp dwell time. This is supported by observations that contiguous adenomas are less prevalent among young adults (34). Although, we cannot exclude the possibility that increases were driven by more efficacious colonoscopy among familial cases, similar increases in adenocarcinoma in a polyp were reported among 20-49-year-olds in USA after excluding familial cases (35). An increase due to enhanced detection is also unlikely because screening is not offered to most adults below age 50 and pre-diagnostic colonoscopy is uncommon in early-onset cases (roughly 6%) (36).

Higher frequency of mucinous or signet ring cell differentiation among early-onset colorectal adenocarcinomas is well-documented (3,17,37-40). This has fuelled conjecture that these are hallmarks of the rising incidence of early-onset colorectal cancer, and that causes for the increase could be causes of mucinous and signet ring cell differentiation (37-40). Our findings challenge this conjecture by demonstrating that i) rates of early-onset colorectal mucinous adenocarcinoma have stabilised or declined since 1990 and ii) both younger and older adults exhibit similar increases in colonic signet ring cell carcinoma. The former observation is also reported in Norway (3) and Canada (30,31). This suggests that causes of the rising incidence are unlikely to be the same as causes of early-onset colorectal cancer *per se*. Increases in signet ring cell carcinoma of the colon, but not rectum, are also reported in South Korea (41), while stable or declining rates are reported in USA (42,43). Reasons behind these trends remain uncertain but warrant further investigation.

Our findings also challenge the notion that the rise in early-onset colorectal cancer is driven by distal colorectal cancers (37-40). Data from Norway (3), Germany (33), and North America (30,31,44) indicate that only distal colorectal cancer is increasing among adults under 50, whereas proximal colon cancer trends are significantly weaker or stable. In contrast, data from Australia (44) and Sweden (45) suggest increases of similar magnitude across distal and proximal colorectal cancers among young adults, consistent with our findings. Since these geographical discrepancies are observed in studies with homogenous site classifications (44,45), one could postulate differing prevalence of risk factors that indiscriminately promote colorectal carcinogenesis. Our observed increases in proximal colon cancer among adults above the screening age (≥75 years) has also been reported in USA (46) and Europe (3,45,47), potentially reflecting higher predisposition of the proximal colon to developing serrated lesions harder to detect by screening (48,49). Nevertheless, our joinpoint modelling identified recent reverses, consistent with delayed effects of screening.

Our study illustrates the importance of stratifying by appendiceal involvement, given the heterogeneity in incidence. After excluding appendix, our estimates of early-onset colon cancer rates are smaller than previous studies in Australia that classified appendix as colon (4). Within appendiceal cancer, there is heterogeneity in the frequency and clinical behaviour of histotypes, from common, mostly indolent neuroendocrine neoplasms in young adults, to advanced older-onset adenocarcinomas and rare but aggressive signet ring cell carcinomas. We found that all these subtypes are increasing at magnitudes higher than any other anatomical site. This may reflect a global phenomenon, with the same observations reported in North America (43,50,51) and Europe (33,52-54).

Whether the rise in appendiceal cancer represents an aetiological phenomenon or artefact of changing diagnostic practices is unknown. Appendiceal tumours are typically incidental discoveries in around 2% of appendectomy specimens for suspected appendicitis (55,56). From 2000-2013, Australia doubled availability to computerised tomography scans (57), and rates of appendectomy rose by 25% (58), which may have fuelled overdiagnosis. Although we could not assess this, increasing appendiceal cancer rates are reported for all stages in North America, after accounting for appendectomies (50). Accordingly, the increase is unlikely to be driven by colonoscopy, which can only inspect the appendiceal orifice and, therefore, lacks specificity for appendiceal cancer (59). Well-differentiated appendiceal neuroendocrine neoplasms were reclassified as malignant by cancer registries around 2015 (60); however, since rates were increasing from 1990, an aetiological explanation seems plausible. One hypothesis is a shift toward managing appendicitis with antibiotics instead of appendectomy, which may promote carcinogenesis via chronic inflammation (61-64).

Neuroendocrine neoplasms exhibited steep increases within the appendix, colorectum, and anus across all ages, and their inclusion inflated estimates of appendiceal and rectal cancer trends. Therefore, increases in early-onset colorectal cancer reported by previous studies that pooled histological subtypes were likely driven, in part, by neuroendocrine neoplasms (2,4). This aligns with previous findings in USA (19), Germany (33), and Norway (3), although neuroendocrine neoplasm trends in these studies were exclusive to appendix and rectum. However, we also detected an increase in colonic neuroendocrine neoplasms, a pattern also reported in Canada (31). Given the absence of age-specific patterning, the increase is unlikely to reflect overdiagnosis: colorectal neuroendocrine neoplasms are typically incidental discoveries on colonoscopy (3,19,32); therefore, one would expect the rise to be exclusive to older populations if driven by screening alone. Rather, the rise may reflect changing classification. In 2000/2004, the World Health Organisation introduced a new classification system for gastrointestinal neuroendocrine neoplasms, and the 2010 edition resulted in most carcinoids being upgraded to grade 1 neuroendocrine neoplasms (65). This may have artefactually inflated rates, although grade information was not available to assess this. Moreover, stage information was not available to assess overdiagnosis, although a study in Norway found increasing rates of colorectal neuroendocrine neoplasms across all stages (3).

The rising incidence of anal squamous cell carcinoma but not adenocarcinoma mirrors trends in high-income countries, following an inverse U-shaped distribution peaking around middle age (21). Although site misclassification cannot be ruled out, the similar trajectory for rectal squamous cell carcinoma might indicate shared aetiology. Human papillomavirus (HPV) is present in 80-90% of anorectal squamous cell carcinomas, but few adenocarcinomas (66,67). Over the last 20 years, incidence of other HPV-related anogenital cancers has increased, and risk factors for anorectal HPV infection have become more prevalent, including lower mean age at first intercourse, greater number of sexual partners, and increasing prevalence of receptive anal intercourse (68-70).

The generational shift in gastrointestinal cancer risk, suggested by birth cohort trends in the present study, is consistent with earlier exposure to risk factors among generations born after 1960. Such factors are likely to selectively increase susceptibility to tumour subtypes increasing in incidence. Sex-related factors may not be critical, as we and others found no prominent difference in trends between men and women (2,4). Inflammatory bowel disease has been implicated, given associations with early-onset colorectal cancer (11) and increasing prevalence since 1970 (71). Diabetes, metabolic syndrome, and childhood obesity are other factors that may explain observed trends, because they have increased in Australia since 1980 and are associated with almost all cancer subtypes exhibiting increasing incidence (10,72,73). Physical inactivity also increased in Australia and is a risk factor for most cancers under study (9,11). Regular aspirin intake reduces colorectal cancer risk after a 10-year lag (74); however, aspirin use in childhood was discouraged in the 1980s due to its association with Reye’s syndrome (75), which may have contributed to the rising early-onset colorectal cancer burden. Although overall alcohol consumption has declined, more Australian’s are undertaking harmful consumption that promotes gastrointestinal carcinogenesis (5,11). Fast food consumption has also increased among young populations since 1970 and has been linked to multiple gastrointestinal cancers (11,76). Meanwhile, protective effects of fruit may have become less apparent given decreasing consumption in Australia over time (6-8).

The effect of other suspected early-life risk factors, and trends in their prevalence, remains poorly un involvement of an altered gut microbiome, supported by changes to dietary patterns and increased antibiotic use in childhood (77). Evidence from animal models and prospective cohorts has linked specific microbes to gastrointestinal carcinogenesis (78,79), and a dose-dependent response between early-life antibiotic use and colorectal cancer risk is suggested (80). Perfluoroalkyl and polyfluoroalkyl substances (PFAS) are man-made fluorinated chemicals widely used in consumer products since 1950, a clear inflection point for birth cohort trends in the present study (81). PFAS are present at unsafe levels in tap water across Australia and have been linked to various gastrointestinal cancers in ecological studies (81). Other suspected early-onset colorectal cancer risk factors based on circumstantial or case-control evidence include bottle feeding, caesarean sections, sulphur microbial diet, vitamin A, and maternal obesity (37-40). To unpack the aetiology moving forward, lifecourse studies are required to understand birth cohort-specific risk factor exposures and age-specific trajectories of susceptibility to tumour subtypes.

Our findings have several public health implications. Increasing incidence in younger generations foreshadow future disease burden as young cohorts carry elevated risk into older age, when cancers most frequently occur. In future, this may tip the benefit-to-harm ratio of screening younger people, which is not currently justified due to low yield. Thus, improving awareness about risk factors, signs, and symptoms remains the primary opportunity for prevention and early detection. Moreover, not only does the rising incidence of squamous cell carcinoma, signet ring cell carcinoma, and neuroendocrine neoplasms dictate future management, since management differs from conventional adenocarcinoma, but it also has implications for screening as incidence continues to rise across young and older groups.

The strengths of this study include nationally representative and complete longitudinal cancer records; systematic evaluation of multiple anatomical sites, including separation of appendiceal and proximal colon cancers; stratification by histological subtype; and inclusion of data from before and after the introduction of population-based screening. In addition, birth cohort analyses provide more robust assessment of trends that may be related to changing risk factor prevalence. Collectively, this enabled detection of age-related differences not previously apparent.

Several limitations should be considered. Individual-level risk factors are not captured in cancer registries; therefore, the impact of these factors on trends could not be quantified. Moreover, we could not disaggregate the impact of changing diagnostic practices, and we lacked information about clinical contexts surrounding diagnosis. Assessing trends by stage and detection method (symptomatic, screening, incidental) could help disentangle the effect of changing generational risk versus diagnostic scrutiny. Additionally, aggregate country-level analyses could mask disparate trends across sociodemographic groups, necessitating further studies to identify high-risk subpopulations. It was also not possible to discern the contribution of hereditary predisposition, although this would unlikely impact trends since most (∼80%) early-onset cases are sporadic (37-40), and prevalence of hereditary syndromes was conceivably stable. Lastly, small case numbers for rare histological subtypes yielded unstable estimates, necessitating caution when interpreting results.

In conclusion, early-onset appendiceal, colorectal, and anal cancer incidence is rising in Australia with variation across age and histology. Overall, the largest number of excess early-onset cases occur in the colorectum. Conventional colorectal adenocarcinoma rates have declined or stabilised in adults over age 50 but continue to rise in adults under 50, potentially due to opposing influences of risk factors and population screening, although this pattern is not observed for other histological subtypes. Anal and rectal squamous cell carcinomas exhibit similar increases that peak around middle age, likely due to HPV infection. Across all ages, increases are most pronounced for neuroendocrine neoplasms and appendiceal cancer irrespective of histology, although the role of changing diagnostic practices and risk factor exposure remains to be determined. Each successive generation born during the second half of the 20^th^ century experienced elevated risk of lower gastrointestinal tumour subtypes with potential distinct and overlapping aetiologies, highlighting the importance of stratifying future studies by site and histology. Extensive efforts are needed to identify factors responsible for these trends to inform prevention strategies.

## Data Availability

National population figures can be extracted via the Australian Bureau of Statistics website (https://www.abs.gov.au/statistics/people/population). All other study data and related documents are available upon request from the corresponding author, Professor Mark Jenkins, via email at, with support from the Australian Institute of Health and Welfare.

## Contributions

**A.L. Meyers:** Conceptualization, Methodology, Software, Formal analysis, Resources, Data Curation, Writing - Original Draft, Writing - Review & Editing; **J.G. Dowty:** Supervision, Writing - Review & Editing; **K. Mahmood:** Supervision, Writing - Review & Editing; **F.A. Macrae:** Methodology, Writing - Review & Editing; **C. Rosty:** Methodology, Writing - Review & Editing; **D.D. Buchanan:** Conceptualization, Methodology, Supervision, Writing - Review & Editing; **M.A. Jenkins:** Conceptualization, Methodology, Resources, Supervision, Writing - Review & Editing. All authors had full access to all study data and had final responsibility for the decision to submit for publication. **A.L. Meyers** and **M.A. Jenkins** have accessed and verified the data.

## Acknowledgements

This work was funded by an Australian Government Research Training Program Scholarship, as well as WP Greene and Rowden White Scholarships from the University of Melbourne (to ALM). DDB is supported by a National Health and Medical Research Council of Australia (NHMRC) Investigator grant (GNT1194896), a University of Melbourne Dame Kate Campbell Fellowship, and by funding awarded to The Colon Cancer Family Registry (CCFR, www.coloncfr.org) from the National Cancer Institute (NCI), National Institutes of Health (NIH) [award U01 CA167551]. MAJ is supported by an NHMRC Investigator grant (GNT1195099), a University of Melbourne Dame Kate Campbell Fellowship, and by funding awarded to the CCFR from NCI, NIH [award U01 CA167551]. The funding sources had no role in study design, data collection, data analysis, data interpretation, or writing of the report. The authors thank the Australian Institute of Health and Welfare and the population-based cancer registries of New South Wales, Victoria, Queensland, Western Australia, South Australia, Tasmania, the Australian Capital Territory and the Northern Territory for the provision of data from the Australian Cancer Database.

## References

1. Bray F, Laversanne M, Sung H, Ferlay J, Siegel RL, Soerjomataram I, et al. Global cancer statistics 2022: GLOBOCAN estimates of incidence and mortality worldwide for 36 cancers in 185 countries. CA Cancer J Clin. 2024;74(1):229–63.

2. Sung H, Siegel RL, Laversanne M, Jiang C, Morgan E, Zahwe M, et al. Colorectal cancer incidence trends in younger versus older adults: an analysis of population-based cancer registry data. Lancet Oncol. 2025;26(1):51–63.

3. Ystgaard MF, Myklebust TÅ, Smeby J, Larsen IK, Guren TK, Kure EH, et al. Early-onset colorectal cancer incidence in Norway: a national registry-based study (1993-2022) analyzing subsite and morphology trends. ESMO Gastrointestinal Oncology. 2025;7(1):100065.

4. Feletto E, Yu XQ, Lew JB, St John DJ, Jenkins MA, Macrae FA, et al. Trends in colon and rectal cancer incidence in Australia from 1982 to 2014: analysis of data on over 375,000 cases. Cancer Epidemiol Biomarkers Prev. 2019;28(1):83–90.

5. Australian Institute of Health and Welfare. Australia’s health 2016. Canberra: AIHW; 2016. Australia’s health series No.: 15. Cat. No.: AUS 199.

6. Walls HL, Magliano DJ, Stevenson CE, Backholer K, Mannan HR, Shaw JE, et al. Projected progression of the prevalence of obesity in Australia. Obesity. 2012;20(1):872–8.

7. Australian Institute of Health and Welfare. A picture of overweight and obesity in Australia. Canberra: AIHW; 2017. Cat. No.: PHE 216.

8. Garnett SP, Baur LA, Jones AMD, Hardy LL. Trends in the prevalence of morbid and severe obesity in Australian children aged 7–15 years, 1985–2012. PLoS One. 2016;11(1):e0154879.

9. Bennie JA, Pedisic Z, van Uffelen JGZ, Gale J, Banting LK, Vergeer I, et al. The descriptive epidemiology of total physical activity, muscle-strengthening exercises and sedentary behaviour among Australian adults – results from the National Nutrition and Physical Activity Survey. BMC Public Health. 2016;16(1):73.

10. Zhou B, Rayner AW, Gregg EW, Sheffer KE, Carrillo-Larco RM, Bennett JE, et al. Worldwide trends in diabetes prevalence and treatment from 1990 to 2022: a pooled analysis of 1108 population-representative studies with 141 million participants. Lancet. 2024;404(10467):2077–93.

11. O’Sullivan DE, Sutherland RL, Town S, Chow K, Fan J, Forbes N, Heitman SJ, et al. Risk factors for early-onset colorectal cancer: a systematic review and meta-analysis. Clin Gastroenterol Hepatol. 2022;20(6):1229–40.

12. Xiao W, Huang J, Zhao C, Ding L, Wang X, Wu B. Diabetes and risks of right-sided and left-sided colon cancer: a meta-analysis of prospective cohorts. Front Oncol. 2022;12(1):737330.

13. World Cancer Research Fund/American Institute for Cancer Research. Continuous update project report. Diet, nutrition, physical activity, and colorectal cancer. London: WCRF; 2017.

14. Robsahm TE, Aagnes B, Hjartaker A, Langseth H, Bray FI, Larsen IK. Body mass index, physical activity, and colorectal cancer by anatomical subsites: a systematic review and meta-analysis of cohort studies. Eur J Cancer Prev. 2013;22(1):492–505.

15. Missiaglia E, Jacobs B, D’ario G, Di Narzo AF, Soneson C, Budinska E, et al. Distal and proximal colon cancers differ in terms of molecular, pathological, and clinical features. Ann Oncol. 2014;25(10):1995–2001.

16. An Y, Zhou J, Lin G, Wu H, Cong L, Li Y, et al. Clinicopathological and molecular characteristics of colorectal signet ring cell carcinoma: a review. Pathol Oncol Res. 2021;27(1):1609859.

17. Collaborative RE, Zaborowski AM, Abdile A, Adamina M, Aigner F, d’Allens L, et al. Characteristics of early-onset vs late-onset colorectal cancer: a review. JAMA Surg. 2021;156(9):865–74.

18. Siegel RL, Torre LA, Soerjomataram I, Hayes RB, Bray F, Weber TK, et al. Global patterns and trends in colorectal cancer incidence in young adults. Gut. 2019;68(12):2179–85.

19. Yao JC, Hassan M, Phan A, Dagohoy C, Leary C, Mares JE, et al. One hundred years after “carcinoid”: epidemiology of and prognostic factors for neuroendocrine tumors in 35,825 cases in the United States. J Clin Oncol. 2008;26(18):3063–72.

20. Bleyer A, Ries LA, Cameron DB, Mansfield SA, Siegel SE, Barr RD. Colon, colorectal and all cancer incidence increase in the Young due to appendix reclassification. JNCI. 2025;djaf038.

21. Kang YJ, Smith M, Canfell K. Anal cancer in high-income countries: increasing burden of disease. PLoS One. 2018;13(10):e0205105.

22. Australian Institute of Health and Welfare. Cancer data in Australia. Canberra: AIHW; 2024.

23. Australian Bureau of Statistics. 3101.0 - Australian historical population statistics, 2020. Canberra: ABS; 2020.

24. Kim HJ, Fay MP, Feuer EJ, Midthune DN. Permutation tests for joinpoint regression with applications to cancer rates. Stat Med. 2000;19(3):335–5.

25. Kim HJ, Chen HS, Midthune D, Wheeler B, Buckman DW, Green D, et al. Data-driven choice of a model selection method in joinpoint regression. J Appl Stat. 2023;50(9):1992–2013.

26. Rosenberg PS, Check DP, Anderson WF. A web tool for age–period–cohort analysis of cancer incidence and mortality rates. Cancer Epidemiol Biomarkers Prev. 2014;23(11):2296–302.

27. Msemburi W, Karlinsky A, Knutson V, Aleshin-Guendel S, Chatterji S, Wakefield J. The WHO estimates of excess mortality associated with the COVID-19 pandemic. Nature. 2023;613(7942):130–7.

28. Joinpoint Regression Program, Version 5.4.0.0 - April 2025; Statistical Methodology and Applications Branch, Surveillance Research Program, National Cancer Institute.

29. R Core Team (2021). R: A language and environment for statistical computing. R Foundation for Statistical Computing, Vienna, Austria. URL https://www.R-project.org/.

30. O’Sullivan DE, Ruan Y, Sutherland RL, Cheung WY, Heitman SJ, Hilsden R, et al. Detailed incidence and mortality trends of early-onset colorectal cancer in Canada. Curr Oncol. 2024;31(12):7765–9.

31. O’Sullivan DE, Ruan Y, Cheung WY, Forbes N, Heitman SJ, Hilsden RJ, et al. Early-onset colorectal cancer incidence, staging, and mortality in Canada: implications for population-based screening. Am J Gastroenterol. 2022;117(9):1502–7.

32. Montminy EM, Zhou M, Maniscalco L, Abualkhair W, Kim MK, Siegel RL, et al. Contributions of adenocarcinoma and carcinoid tumors to early-onset colorectal cancer incidence rates in the United States. Ann Intern Med. 2021;174(2):157–66.

33. Voigtländer S, Hakimhashemi A, Grundmann N, Rees F, Meyer M, Algül H, et al. Trends of colorectal cancer incidence according to age, anatomic site, and histological subgroup in Bavaria: a registry-based study. Front Oncol. 2022;12(1):904546.

34. Merrill RM, Anderson AE. Risk-adjusted colon and rectal cancer incidence rates in the United States. Dis Colon Rectum. 2011;54(10):1301–6.

35. Aloysius MM, Nikumbh T, Yadukumar L, Asija U, Shah NJ, Aswath G, et al. National trends in the incidence of sporadic malignant colorectal polyps in young patients (20-49 Years): an 18-year SEER database analysis. Medicina. 2024;60(4):673.

36. Montminy EM, Zhou M, Maniscalco L, Penrose H, Yen T, Patel SG, et al. Trends in the incidence of early-onset colorectal adenocarcinoma among black and white US residents aged 40 to 49 years, 2000-2017. JAMA Netw Open. 2021;4(11):e2130433.

37. Young JP, Win AK, Rosty C, Flight I, Roder D, Young GP, et al. Rising incidence of early-onset colorectal cancer in A ustralia over two decades: report and review. J Gastroenterol Hepatol. 2015;30(1):6–13.

38. Patel SG, Karlitz JJ, Yen T, Lieu CH, Boland CR. The rising tide of early-onset colorectal cancer: a comprehensive review of epidemiology, clinical features, biology, risk factors, prevention, and early detection. Lancet Gastroenterol Hepatol. 2022;7(3):262–74.

39. Garrett C, Steffens D, Solomon M, Koh C. Early-onset colorectal cancer: why it should be high on our list of differentials. ANZ J Surg. 2022;92(7-8):1638-43.

40. Sinicrope FA. Increasing incidence of early-onset colorectal cancer. N Engl J Med. 2022;386(16):1547–58.

41. Kim JH, Kim H, Kim JW, Kim HM. Trends in the incidence and survival rates of colorectal signet-ring cell carcinoma in the South Korean population: analysis of the Korea Central Cancer Registry Database. J Clin Med. 2021;10(18):4258.

42. Mehmood T. Comparison of incidence and survival outcomes in mucinous and signet-ring cell colorectal cancers with classical adenocarcinoma: a SEER analysis. Ann Oncol. 2018;29(1):39.

43. Li H, Zong Z, Zhou T, Sun L, Wang A, Zhang K, et al. Trends of incidence and survival in patients with gastroenteropancreatic signet ring cell carcinoma: an analysis from the Surveillance, Epidemiology, and End Results program. J Gastrointest Oncol. 2019 Oct;10(5):979.

44. Araghi M, Soerjomataram I, Bardot A, Ferlay J, Cabasag CJ, Morrison DS, et al. Changes in colorectal cancer incidence in seven high-income countries: a population-based study. Lancet Gastroenterol Hepatol. 2019;4(7):511–8.

45. Gutlic I, Schyman T, Lydrup ML, Buchwald P. Increasing colorectal cancer incidence in individuals aged < 50 years—a population-based study. Int J Colorectal Dis. 2019;34(1):1221–6.

46. Tadros M, Mago S, Miller D, Ungemack JA, Anderson JC, Swede H. The rise of proximal colorectal cancer: a trend analysis of subsite specific primary colorectal cancer in the SEER database. Ann Gastroenterol. 2021;34(4):559.

47. Granger SP, Preece RA, Thomas MG, Dixon SW, Chambers AC, Messenger DE. Colorectal cancer incidence trends by tumour location among adults of screening-age in England: a population-based study. Colorectal Dis. 2023;25(9):1771–82.

48. Hoffmeister M, Bläker H, Jansen L, Alwers E, Amitay EL, Carr PR, et al. Colonoscopy and reduction of colorectal cancer risk by molecular tumor subtypes: a population-based case-control study. Am J Gastroenterol. 2020;115(12):2007–16.

49. Chang LC, Shun CT, Hsu WF, Tu CH, Tsai PY, Lin BR, et al. Fecal immunochemical test detects sessile serrated adenomas and polyps with a low level of sensitivity. Clin Gastroenterol Hepatol. 2017;15(6):872–9.

50. Singh H, Koomson AS, Decker KM, Park J, Demers AA. Continued increasing incidence of malignant appendiceal tumors in Canada and the United States: a population-based study. Cancer. 2020;126(10):2206–16.

51. Marmor S, Portschy PR, Tuttle TM, Virnig BA. The rise in appendiceal cancer incidence: 2000-2009. J Gastrointest Surg. 2015;19(4):743–50.

52. Orchard P, Preece R, Thomas MG, Dixon SW, Wong NA, Chambers AC, et al. Demographic trends in the incidence of malignant appendiceal tumours in England between 1995 and 2016: population-based analysis. BJS Open. 2022;6(4):zrac103.

53. Van Den Heuvel MG, Lemmens VE, Verhoeven RH, De Hingh IH. The incidence of mucinous appendiceal malignancies: a population-based study. Int J Colorectal Dis. 2013;28(1):1307–10.

54. Johansson J, Andersson RE, Landerholm K, Redéen S. Incidence of appendiceal malignancies in Sweden between 1970 and 2012. World J Surg. 2022;44(12):4086–92.

55. Leonards LM, Pahwa A, Patel MK, Petersen J, Nguyen MJ, Jude CM. Neoplasms of the appendix: pictorial review with clinical and pathologic correlation. Radiographics. 2017;37(4):1059–83.

56. Kohler F, Rosenfeldt M, Matthes N, Kastner C, Germer CT, Wiegering A. Incidental finding of mucinous neoplasia of the appendix: treatment strategies. Chirurg. 2019;90(3):194–201.

57. Senate Standing Committees on Community Affairs. Availability and accessibility of diagnostic imaging equipment around Australia: the senate community affairs committee secretariat. Canberra: APH; 2018.

58. Australian Commission on Safety and Quality in Health Care. The Second Australian Atlas of Healthcare Variation. Sydney: ACSQHC; 2017.

59. Trivedi AN, Levine EA, Mishra G. Adenocarcinoma of the appendix is rarely detected by colonoscopy. J Gastrointest Surg. 2009;13(1):668–75.

60. Siegel RL, Miller KD, Goding Sauer A, Fedewa SA, Butterly LF, Anderson JC, et al. Colorectal cancer statistics, 2020. CA Cancer J Clin. 2020;70(3):145–64.

61. Brunner M, Lapins P, Langheinrich M, Baecker J, Krautz C, Kersting S, et al. Risk factors for appendiceal neoplasm and malignancy among patients with acute appendicitis. Int J Colorectal Dis. 2020;35(1):157–63.

62. Loftus TJ, Raymond SL, Sarosi Jr GA, Croft CA, Smith RS, Efron PA, et al. Predicting appendiceal tumors among patients with appendicitis. J Trauma Acute Care Surg. 2017;82(4):771–5.

63. Lietzén E, Grönroos JM, Mecklin JP, Leppäniemi A, Nordström P, Rautio T, et al. Appendiceal neoplasm risk associated with complicated acute appendicitis-a population based study. Int J Colorectal Dis. 2019;34(1):39–46.

64. Andersson RE, Petzold MG. Nonsurgical treatment of appendiceal abscess or phlegmon: a systematic review and meta-analysis. Ann Surg. 2007;246(5):741–8.

65. International Agency for Research on Cancer. World Health Organization classification of tumours, pathology and genetics of tumours of the digestive system. Lyon: IARC Press, 2010.

66. Mignozzi S, Santucci C, Malvezzi M, Levi F, La Vecchia C, Negri E. Global trends in anal cancer incidence and mortality. Eur J Cancer Prev. 2024;33(2):77–86.

67. Welten VM, Fields AC, Malizia RA, Yoo J, Irani JL, Bleday R, et al. The association between sex and survival for anal squamous cell carcinoma. J Surg Res. 2021;268(1):474–84.

68. Herbenick D, Reece M, Schick V, Sanders SA, Dodge B, Fortenberry JD. Sexual behavior in the United States: results from a national probability sample of men and women ages 14-94. J Sex Med. 2010;7(Suppl 5):255–65.

69. Herbenick D, Reece M, Schick V, Sanders SA, Dodge B, Fortenberry JD. Sexual behaviors, relationships, and perceived health status among adult women in the United States: results from a national probability sample. J Sex Med. 2010;7(Suppl 5):277–90.

70. McBride KR, Fortenberry JD. Heterosexual anal sexuality and anal sex behaviors: a review. J Sex Res. 2010;47(1):123–36.

71. Alatab S, Sepanlou SG, Ikuta K, Vahedi H, Bisignano C, Safiri S, et al. The global, regional, and national burden of inflammatory bowel disease in 195 countries and territories, 1990-2017: a systematic analysis for the Global Burden of Disease Study 2017. Lancet Gastroenterol Hepatol. 2020;5(1):17–30.

72. Phelps NH, Singleton RK, Zhou B, Heap RA, Mishra A, Bennett JE, et al. Worldwide trends in underweight and obesity from 1990 to 2022: a pooled analysis of 3663 population-representative studies with 222 million children, adolescents, and adults. Lancet. 2024;403(10431):1027–50.

73. Zhou B, Bennett JE, Wickham AP, Singleton RK, Mishra A, Carrillo-Larco RM, et al. General and abdominal adiposity and hypertension in eight world regions: a pooled analysis of 837 population-based studies with 7.5 million participants. Lancet. 2024;404(10455):851–63.

74. Emery JD, Nguyen P, Minshall J, Cummings KL, Walker J. Chemoprevention: a new concept for cancer prevention in primary care. Aust J Gen Pract. 2018;47(12):825–8.

75. Therapeutic Goods Administration. Review of Aspirin/Reye’s Syndrome warning statement. Canberra: TGA; 2004.

76. Guthrie JF, Lin BH, Frazao E. Role of food prepared away from home in the American diet, 1977-78 versus 1994-96: changes and consequences. J Nutr Educ Behav. 2002;34(3):140–50.

77. Allwell-Brown G, Hussain-Alkhateeb L, Kitutu FE, Strömdahl S, Mårtensson A, Johansson EW. Trends in reported antibiotic use among children under 5 years of age with fever, diarrhoea, or cough with fast or difficult breathing across low-income and middle-income countries in 2005-17: a systematic analysis of 132 national surveys from 73 countries. Lancet Glob Health. 2020;8:e799–807.

78. Zepeda-Rivera M, Minot SS, Bouzek H, Wu H, Blanco-Míguez A, Manghi P, et al. A distinct Fusobacterium nucleatum clade dominates the colorectal cancer niche. Nature. 2024;628(8007):424–32.

79. Nguyen LH, Cao Y, Hur J, Mehta RS, Sikavi DR, Wang Y, et al. The sulfur microbial diet is associated with increased risk of early-onset colorectal cancer precursors. Gastroenterology. 2021;161(5):1423–32.

80. Boursi B, Haynes K, Mamtani R, Yang YX. Impact of antibiotic exposure on the risk of colorectal cancer. Pharmacoepidemiol Drug Saf. 2015;24(1):534–42.

81. IARC Working Group on the Identification of Carcinogenic Hazards to Humans. Perfluorooctanoic acid (PFOA) and perfluorooctanesulfonic acid (PFOS). IARC Monographs on the Identification of Carcinogenic Hazards to Humans. Volume 135. Lyon (FR): IARC; 2024.

